# Distinct single cell gene expression in peripheral blood monocytes correlates with tumor necrosis factor inhibitor treatment response groups defined by type I interferon in rheumatoid arthritis

**DOI:** 10.1101/19011460

**Authors:** Theresa L. Wampler Muskardin, Wei Fan, Zhongbo Jin, Mark A. Jensen, Jessica M. Dorschner, Yogita Ghodke-Puranik, Betty Dicke, Danielle Vsetecka, Kerry Wright, Thomas Mason, Scott Persellin, Clement J. Michet, John M. Davis, Eric Matteson, Timothy B. Niewold

## Abstract

Previously, we demonstrated that type I IFN (IFNβ/α) activity can predict non-response to tumor necrosis factor inhibitors (TNFi) in rheumatoid arthritis (RA). In this study, we examine the biology of TNFi non-response in monocytes from RA patients. We compared single cell gene expression in purified classical (CL, n=342) and non-classical (NC, n=359) monocytes. RA patients were grouped according to their pre-TNFi IFNβ/α activity: those likely to have EULAR no response (non-response group, IFNβ/α 0 or >1.3, n=9) and those likely to have EULAR moderate or good response (response group, IFNβ/α > 0 and ≤1.3, n=6). Major differences in gene expression were apparent in principal component and unsupervised cluster analyses. CL monocytes from the non-response group were unlikely to express *JAK1* and *IFI27* (p <0.0001 and p 0.0005, respectively). In NC monocytes from the same group, expression of *IFNAR1, IRF1, TNFA, TLR4* (p ≤0.0001 for each) and others was enriched. Interestingly, *JAK1* expression was absent in CL and NC monocytes from 9 patients. This pattern strongly associated with the non-response IFNβ/α group, suggesting a major biological difference between *JAK1* expressors and non-expressors. The type I IFN activity that was previously found to predict TNFi response associated with changes in gene expression in monocytes that suggest differential IFN pathway activation in RA patients who are either likely to respond or to have no response to TNFi. This work could suggest mechanisms for TNFi non-response, and potential therapeutic strategies for those patients unlikely to respond to TNFi.

## 1 Introduction

Rheumatoid Arthritis (RA) is the most common inflammatory joint disease world-wide, characterized by a destructive arthritis and serious extra-articular manifestations, including accelerated vascular disease(*1*). Early, effective treatment prevents damage. Thus, remission or very low disease activity within the first 3 months is the goal (*2, 3*). New therapies have made remission possible for a greater number of patients. However, the current treatment strategy is one of trial-and-error, as we are not able to predict which medication will work for an individual patient. Tumor necrosis factor inhibitors (TNFi) are the most common biologic treatment employed(*4*). Responses are variable, with approximately 30% not responding and another 30% achieving only partial response. Insufficient treatment is associated with increased morbidity, mortality, and a heavy economic burden (*5-8*). Type I IFN levels are genetically determined to some degree (*9, 10*) and type I IFNs are pleiotropic biologic response modifiers, making them ideal candidate biomarkers for response to immunomodulatory therapies. Recently, we studied pre-treatment circulating IFN-alpha (IFNα), IFN-beta (IFNβ), and total type I IFN activity in RA patients just prior to receiving a TNFi (*11*) in independent test and validation cohorts. The ratio of IFNβ to IFNα activity (IFNβ/α) >1.3 was strongly predictive of non-response to TNFi therapy (specificity=77% in the validation cohort). Remarkably, no patient with a ratio >1.3 achieved remission or low disease activity.

Monocytes are one of the major effector cells in RA (*12, 13*). By studying the effect of IFNβ/α ratio on monocytes, we understand the functional impact of the IFN ratio on a key effector cell type. This may suggest cellular mechanisms that underlie response/non-response to TNFi therapy in RA. We chose to study gene expression in single cells, as effects of IFN on single immune cells or cell types may be masked in whole blood or mixed cell populations (*14*). We find that circulating type I IFN ratio corresponds to strikingly different gene expression patterns in RA patient monocytes, and that particular transcripts such as *JAK1* are highly informative and could suggest alternate therapeutic avenues in patients who are predicted to be TNFi non-responders.

## 2 Material and Methods

### 2.1 Patient and Public Involvement

Patients/the public were not involved in the design of the study. The study design and plans to disseminate study results to participants were informed by patient priorities and preferences.

### 2.2 Patients and Samples

Blood samples from 15 patients with RA were recruited from the Mayo Clinic in Rochester, Minnesota, USA. All of the patients fulfilled the 2010 American College of Rheumatology classification criteria for RA (*15*) and were seropositive. Exclusion criteria included overlap autoimmune connective tissue disease, pregnancy, active acute infection, chronic infection (e.g., hepatitis C, HIV, etc.), current intravenous therapy (e.g., methylprednisolone or cyclophosphamide), and history of biologic therapy. All samples were obtained prior to initiation of TNFi. All patients provided informed consent, and the study was approved by the institutional review board. Subjects were grouped by pre-TNFi serum type I IFN activity into two groups, those with detectable type I IFN activity but low IFNβ/α ratio (IFNβ/α > 0 and ≤1.3, n=6, responder group), and those with either undetectable type I IFN activity or a high IFNβ/α ratio (IFNβ/α 0 or >1.3, n=9). Patients with undetectable type I IFN activity typically did not respond to TNFi therapy (*11*), and so these patients were grouped together with those who have an IFNβ/α ratio >1.3 (non-responder group).

### 2.3 Determination of IFNβ/α ratio

Type I IFN activity in serum was measured using a validated functional assay in which reporter cells are used to measure the ability of patient sera to cause type I IFN-induced gene expression (*16*). Reporter cells (WISH cells, ATCC #CCL-25) were cultured with patient serum for 6 hours. Cells were then lysed, and cDNA was made from total cellular mRNA. Canonical type I IFN-induced gene expression (*MX1, PKR*, and *IFIT1*) (*17*), was measured using qPCR. The relative expression of these three genes was standardized to healthy donors and summed to generate a score reflecting the ability of sera to cause type I IFN-induced gene expression (serum type I IFN activity). This assay has been informative in a wide range of autoimmune diseases (*11, 16, 18-20*), and we have not found significant functional inhibitors in samples studied to date (*21*). Additional aliquots were tested following pre-incubation with polyclonal anti-IFNα (19.6 μg/mL, PBL Assay Science) and anti-IFNβ (10.1 μg/mL, Chemicon) antibodies. The amount of inhibition of the observed type I IFN activity by anti-IFNα antibody allowed for quantitative assessment of IFNβ activity, and that by antiβ antibody allowed for quantitative assessment of IFNα activity. The ratio of IFNβ activity to IFNα activity (IFNβ/α activity ratio) was then calculated for each serum sample using these data. Those samples reading very low (<1 pg/mL) for total type I IFN activity were categorized as not having significant type I IFN present, and no ratio was calculated.

### 2.4 Purification of classical (CD14^++^CD16^-^) and non-classical (CD14^dim^CD16^+^) monocytes

Classical (CL) and non-classical (NC) monocytes were isolated from peripheral blood using the protocol described in Jin et al(*22*). Briefly, CL (CD14++CD16−) monocytes were purified using the Human Pan-Monocyte Isolation Kit (Miltenyi) with modification of adding anti-CD16-biotin (Miltenyi) into the biotin–antibody cocktail. CD14+ selection (Miltenyi) was used subsequently to further increase purity. Purified CL monocytes were stained with Molecular Probes CellTracker Green CMFDA Dye (Life Technologies). NC (CD14^dim^CD16+) monocytes were purified similarly, using CD16 microbeads (Miltenyi) during positive selection. Purity checked by flow cytometry was very high (> 95%) for both CL and NC monocytes.

### 2.5 C1 single cell isolation and measurement of gene expression

Using the Fluidigm C1 Single-Cell Auto Prep System we isolated single cells from the bulk monocyte subsets. NC and CL monocytes were loaded onto the C1 Integrated Fluidic Circuit (IFC) sequentially. Determination of NC or CL lineage of individual cells was made by direct visualization using fluorescent microscopy. Pre-amplification was done using the C1 Single-Cell Auto Prep Array IFC, according to the manufacturer’s protocol. Melt curves were inspected to ensure that all PCR products were uniform. Amplification curves were analyzed and those not following the expected log-growth curve were excluded. 87 target genes were analyzed (Supplemental Table 1). Target gene pre-amplified cDNAs were assayed using 96.96 IFCs on the BioMark HD System (Fluidigm) according to the manufacturer’s protocol. Empty wells and wells that contained more than one cell after C1 automated single cell capture were identified by visual inspection using microscopy. These wells were excluded from the dataset. A cell was also removed from the dataset if failure score (total CT value) was greater than two standard deviations above the mean, as this indicated that the cell’s overall expression level was too low to be trusted for downstream analysis.

**Table 1.** General characteristics, disease activity measures, and medications of RA patients.

| Characteristic | IFN-$\beta/\alpha$ 0 *or* > 1.3 (n=9) | IFN-$\beta/\alpha$ > 0 *and* ≤ 1.3 (n=6) |
| --- | --- | --- |
| Age (mean,range)* | 54 (27 - 75) | 55 (42 - 70) |
| Gender (F, M) | 4F, 5M | 4F, 2M |
| CCP positivity (n, %) | 8 (89) | 6 (100) |
| RF positivity (n, %) | 7 (78) | 5 (83) |
| ESR (median, range) | 7.5 (0 - 41) | 21.5 (13 - 40) |
| DAS28-CRP (mean ± std.dev, range)* | 2.92 ± 1.82<br>1.15 - 5.77 | 3.42 ± 0.62<br>2.52 - 4.23 |
| Medications (n, %) |  |  |
| Prednisone | 4 (44) | 1 (17) |
| *median dose (mg/day)* | *10* | *5* |
| *dose range (mg/day)* | *5 - 15* | *5* |
| NSAIDs (prn) | 0 | 3 (50) |
| Methotrexate | 8 (89) | 3 (50) |
| *median dose (mg/wk)* | *20* | *10* |
| *dose range (mg/wk)* | *17.5 - 25* | *7.5 - 20* |
| Sulfasalazine | 2 (22) | 0 |
| Leflunomide | 2 (22) | 0 |
| Hydroxychloroquine | 5 (55) | 2 (33) |
| Statin | 3 (33) | 1 (17) |
| ASA-81 | 2 (22) | 0 |
| Allopurinol | 1 (11) | 0 |

### 2.6 Statistical Analysis

Principal component analysis was used to reduce dimensionality in the complex data sets, and compare overall trends between patient groups and CL and NC monocytes. Unsupervised hierarchical clustering was done to detect individual genes and gene sets that defined the patient groups and to identify other possible strata within the data. Each gene was also tested individually for association with patient group, using Mann Whitney U testing for the quantitative data and Fisher’s exact test for the categorical expressed/not expressed analysis. For these analyses, we used the following strategy to account for multiple comparisons. We expected to find correlations between transcripts in the same cell, which would make a strict Bonferroni correction inappropriate, as each of the 87 tests is not independent. So we first calculated pairwise Spearman correlations (rho) for each possible pairing of transcripts, resulting in 3741 pairwise correlations. The average correlation between transcripts was then calculated and a threshold p-value was derived using the following modified Bonferroni method to account for between-transcript correlations:

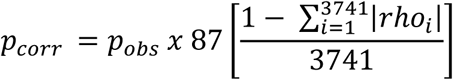

This resulted in a threshold p-value of <0.0008 for a corrected alpha of 0.05.

## 3 Results

### 3.1 Circulating type I IFN ratio corresponds to large differences in monocyte gene expression

Among the participants in the groups, there were no significant differences in age or disease activity score (DAS), and treatments were comparable (Table 1). We were able to analyze results of 87 target genes from 701 individual monocytes (342 CL, 359 NC). Principal component analysis (PCA) showed large differences in the second principal component in CL Mo when cells are labeled by their patient group category. Similar findings were observed in the NC monocytes, although the clustering by patient group related to the second principal component on the PCA plot was not as strong in NC cells as in CL cells (Figure 1). We next performed unsupervised hierarchical clustering of the target genes to visualize the difference between groups with respect to individual transcripts (Figure 2). In this analysis, it was clear that certain genes strongly aligned with the type I IFN activity groups. In particular, *JAK1* appeared to be strongly predictive of patient group (Figure 2).

**Figure 1.**
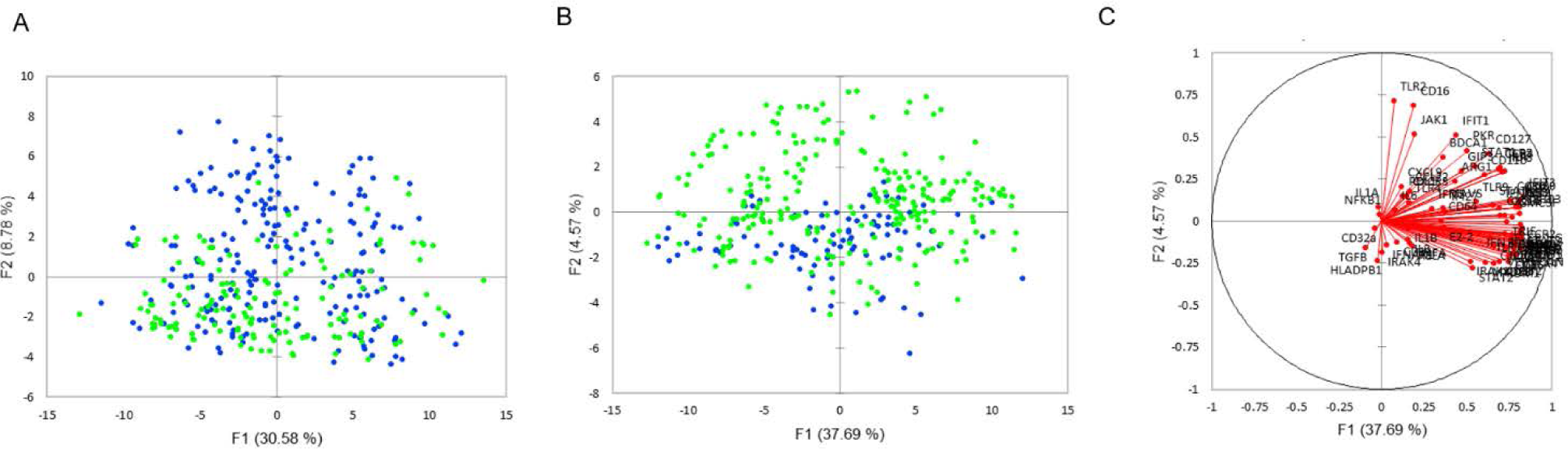
Principal component analysis (PCA) of single monocyte gene expression. Plots depict the first two differentiating factors among the gene expression data of non-classical monocytes (A), and classical monocytes (B). Blue 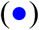 denotes data from subjects in the “IFNβ/α 0 or > 1.3” group. Green 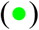 denotes data from subjects in the “IFNβ/α > 0 and ≤ 1.3” group. In B, most of the monocytes from patients in the IFNβ/α 0 or > 1.3 group are on one side of the Y axis; thus, the pretreatment type I IFN-β/α ratio appears to impact classical monocyte gene expression. Active variables (C) for classical monocyte PCA are shown.

**Figure 2.**
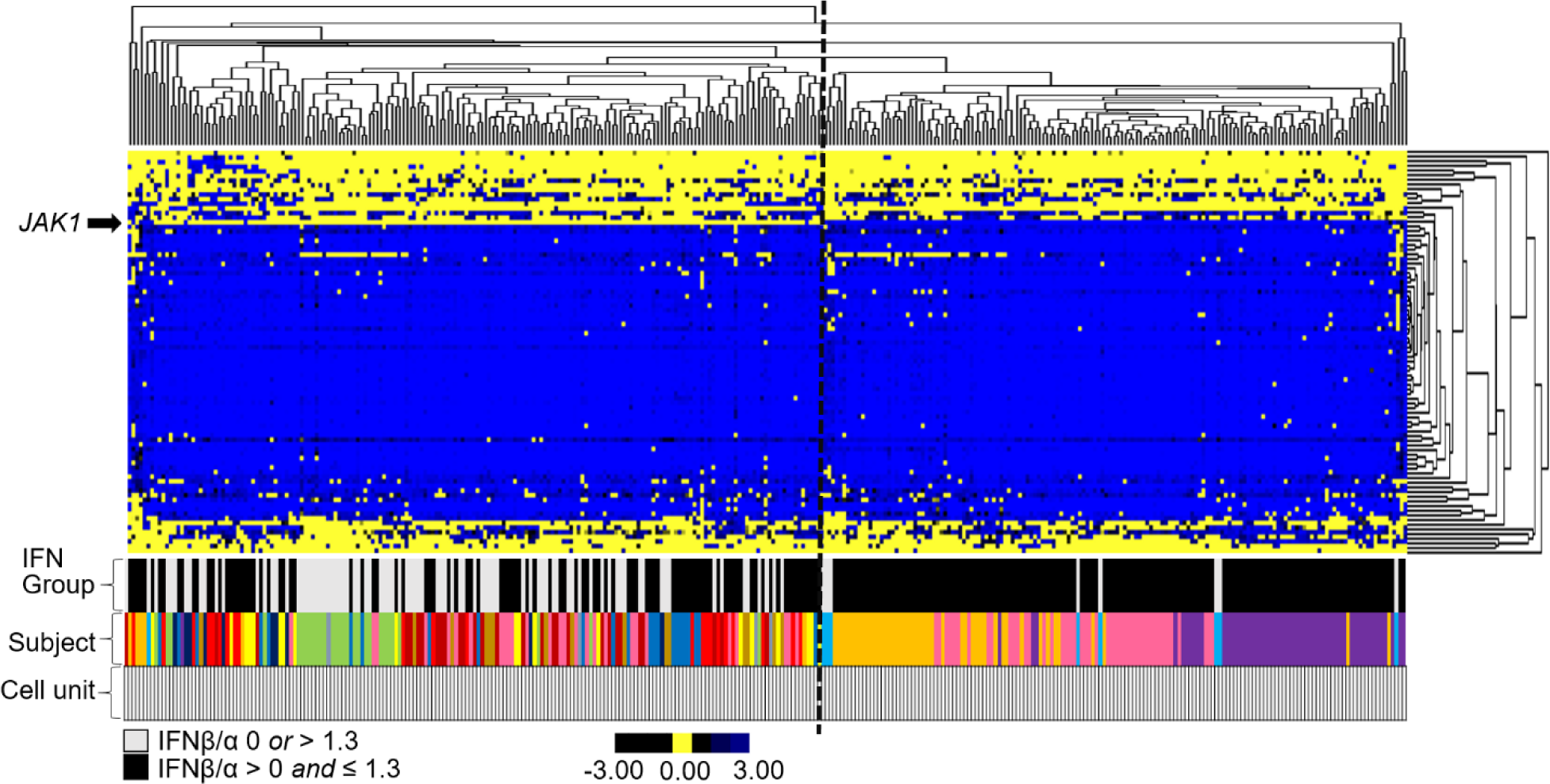
Unsupervised hierarchical clustering of single cell pre-biologic gene expression in classical monoctyes. Genes (Y-axis). Single classical monocytes (X-axis). Both genes and cells were selected for clustering. The bars under the heatmap indicate the IFNβ/α group, subject, and width of data depicted that is from a single cell. Each subject is depicted by a different color. The bottom bar shows the width of data depicted that is from a single cell. See legends for relative expression level (yellow/black/blue) and IFNβ/α group (black/light grey) color assignments.

### 3.2 Different genes were associated with blood IFN ratio in CL vs. NC cells

In the categorical expressed/not expressed analysis, there were significant differences in the transcripts observed between type I IFN activity groups in CL as compared to NC monocytes. In CL monocytes, in addition to JAK1, IFI27 was less likely to be expressed in the non-response IFNβ/α 0 or >1.3 group (Table 2). Thus, the two significant findings in this analysis in CL monocytes were both less likely to be expressed in the non-response group. In NC monocytes, a number of transcripts were more likely to be expressed in the non-response (IFNβ/α 0 or >1.3) group. These include HLADRB1, TNFA, PDL1, TGFB, CD11c, IL8, and IFNAR1 (Table 2). One transcript, JAK1, was less likely to be expressed in the non-response (IFNβ/α 0 or >1.3) group (Table 2).

**Table 2.**
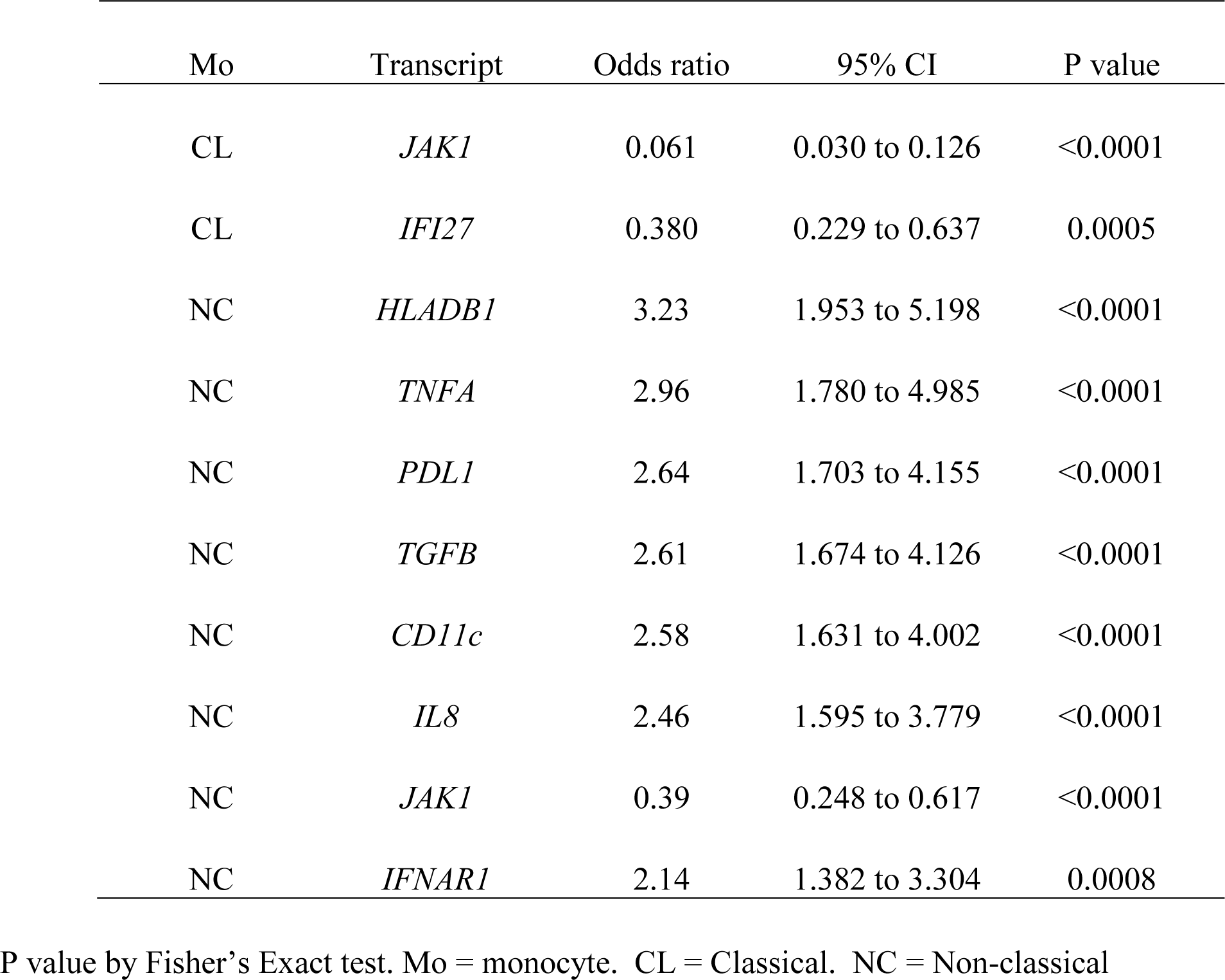
Odds of being expressed in the TNFi non-response (IFNβ/α 0 or > 1.3) group.

Examination of the quantitative data resulted in some similar findings and some additional findings became apparent. In CL monocytes, many genes were reduced in expression in the IFNβ/α 0 or >1.3 group, including JAK1 and IFI27 which were significant in the expressed/not expressed analysis. Additional transcripts that were reduced in the IFNβ/α 0 or >1.3 group in quantitative analysis were: TLR7, TLR8, TLR2, MAVS, PKR, GMCSF, IRF8, IL4, IL1A, ILT7, CD127, CD16, and CCR4 (Supplemental Figure A). In NC Mo, a large number of genes showed increased expression in the IFNβ/α 0 or >1.3 group, including HLADRB1, TNFA, PDL1, TGFB, CD11c, IL8, and IFNAR1 which were identified in the expressed/not expressed analysis. Additional genes that were significantly increased in the in the IFNβ/α 0 or >1.3 group in quantitative analysis were: TLR4, MYD88, IRF1, FCER1G and CD86 (Supplemental Figure B).

Genes that differed between groups by non-parametric (Mann Whitney U) univariate analysis were tested in multivariate logistic regression models. In CL Mo, JAK1, TLR2, IRF8, CD16, and IL1A were retained as independent factors predictive of type I IFN activity group (Table 3).

**Table 3.** Retained transcripts for prediction of patient group by multivariate logistic regression.

| Mo | Transcript | Coefficient | P value |
| --- | --- | --- | --- |
| CL | *JAK1* | -0.35391 | <0.0001 |
| CL | *TLR2* | -0.50455 | 0.0003 |
| CL | *IRF8* | -0.64969 | 0.002 |
| CL | *CD16* | -0.44492 | 0.029 |
| CL | *IL1A* | -0.28175 | 0.035 |
| NC | *CD86* | 0.22951 | 0.001 |
| NC | *HLADRB1* | 0.097105 | 0.016 |
| NC | *IL8* | 0.11685 | 0.002 |
| NC | *PDL1* | 0.099174 | 0.037 |
| NC | *TGFB* | 0.07669 | 0.042 |
| NC | *FCER1G* | -0.25984 | 0.034 |

Mo = monocyte. CL = Classical. NC = Non-classical

In NC Mo, CD86, HLADRB1, IL8, PDL1, TGFB, and FCER1G were retained in the model. (Table 3). ROC curve analysis of these transcripts demonstrated an area under the curve (AUC) of 0.89 (Std. error 0.0172, 95% CI 0.849-0.919) and 0.76 (Std. error 0.026, 95% CI 0.708-0.799), respectively.

### 3.3 JAK1 expression was completely suppressed in some RA patients, and this suppression correlated strongly with non-response (IFNβ/α 0 or >1.3)

*JAK1* was unlikely to be expressed in both CL and NC monocytes from patients in the IFNβ/α 0 or > 1.3 group (Odds 0.06, p value < 0.0001, 95% CI 0.03-0.13 in CL; Odds 0.39, corrected p value < 0.0063, 95% CI 0.25-0.62 in NC). Ninety-one percent of CL monocytes and seventy-six percent of NC monocytes in the IFNβ/α 0 or > 1.3 group did not express JAK1. Whereas in the other IFN group (IFNβ/α > 0 ≤ 1.3), the majority of CL monocytes (63%) and forty-five percent of NC monocytes expressed JAK1. Strikingly, only one participant in the IFNβ/α 0 or > 1.3 group expressed JAK1 in CL monoctyes (Figure 3). Interestingly, after enrollment into our study, this participant was found to have several pre-malignant melanoma lesions that were ultimately removed near the time she began TNFi. Melanocytes and melanoma cells produce IFNβ and are capable of suppressing their own proliferation by secretion of endogenous IFNβ (*23*). It is possible that the increased IFNβ/α activity noted in this participant was significantly influenced by the pre-malignant melanoma and less informative as a physiologic immune phenotype predictive of response to TNFi.

**Figure 3.**
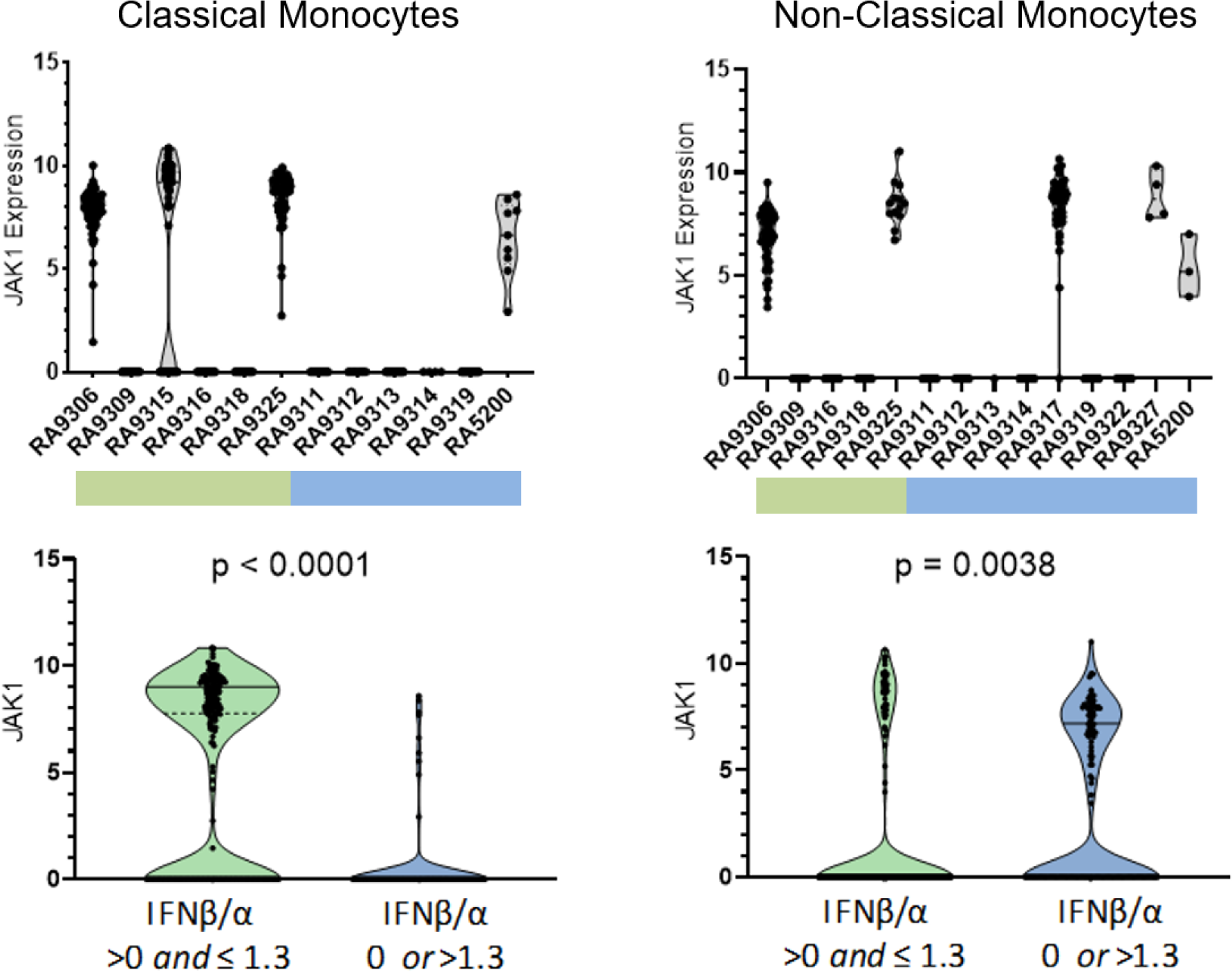
Expression of JAK1 in single classical and non-classical monocytes. P-value by non-parametric Mann Whitney U. Top panels show each individual patient’s cells in a separate column, bottom panels show cells from all patients in aggregate.

The participants who did not express JAK1 in CL monocytes also did not express JAK1 in NC monocytes (Figure 3). This “on or off” expression pattern was not seen in any of the other transcripts studied, and was not observed in healthy controls (data not shown). In the JAK1 expressors, most of the cells expressed JAK1 (83% of cells in CL, 99% in NC). Given this observed distribution, even a subject with only 5 CL cells represented would have an extremely small (0.01%) chance for miscategorization due to sampling error. The same primers were used to measure expression in all experiments, and also have been used to study healthy controls and lupus patients in other studies in which this pattern of JAK1 expression was not observed (22). Each of the 96-well plates were run independently on different days, and it would be highly unlikely that the same one out of the 87 target genes would fail each time, and largely in those patients with a non-response IFN ratio. No other transcript shared this expression pattern in our study.

## 4 Discussion

Using a novel single cell PCR approach, we used a panel of type I IFN and innate immune system related genes to define gene expression states in monocytes from RA patients grouped by their pre-treatment blood type I IFN activity that would predict response or non-response to TNFi. Limitations of this study include the number of patients studied. As is common with single cell gene expression studies, a large number of cells are studied from a more modest number of people. Despite this, we observed fascinating patterns that were shared across different subjects. The technology we used to capture cells does not capture as many cells as some other platforms, such as droplet sequencing. However, in contrast to the droplet sequencing methods which were available at the time of our study, we were able to first isolate and purify CL and NC monocyte subsets *a priori*, instead of defining cell populations afterward using transcriptional patterns to infer lineages. This method also allowed us to intentionally increase the number of NC monocytes examined, which are comparatively rare in circulation and thus less deeply studied by droplet RNA-seq technology. While RNA-seq would provide total transcriptome data, PCR data is typically more robust and more quantitative, and we found that our quantitative analyses both confirmed and extended the findings observed in the expressed/not expressed analyses. Working with this more limited panel of carefully quantitated genes led to novel insights in our study.

We observed striking differences in gene expression patterns in circulating CL and NC monocytes between those subjects likely to respond to TNFi as compared to those that are not likely to respond. Our data suggest differential IFN pathway activation in monocyte subsets from patients who do not respond to TNFi. The outcome of type I IFN receptor activation depends on the pathway components present in the cell and the context (e.g., other cytokines can influence the outcome of IFN receptor ligation) (*24, 25*). Murine data has shown that JAK1 plays an essential and non-redundant role in promoting biologic responses induced by class II cytokine receptor family members, including the receptors for type I IFNs, type II IFN, and IL-10 (*26*). JAK1 is required for canonical type I IFN pathway signaling. In our data, it was striking that JAK1 expression was absent in some patients’ monocytes, and that this was a strong predictor of TNFi response group. The pattern in which none of the monocytes studied in a given subject expressed JAK1 was observed only in patients and not in controls, suggesting that a disease related factor may be contributing to this pattern, such as a cytokine signal inducing a strong transcriptional repressor. It is possible that the type I IFNs could contribute to this process, given that this pattern was associated with IFN ratio in our study. Production of type I IFNs depends on cell type and context (*27-30*). An RA patient’s circulating IFNβ/α activity may reflect different numbers of cells which produce IFNα and IFNβ in the inflamed tissue. Also, steady-state levels of IFNβ can be influenced by our microbiota (*31*), which raises the possibility that circulating IFNβ activity could correlate with components of a patient’s microbiome.

In addition to JAK1, the IFNAR1, IFI27, PKR, and TNFA transcripts were differentially expressed between the response groups. IFNAR1 expression, but not IFNAR2, was enriched in NC Mo of participants in the TNFi non-response (IFNβ/α 0 or >1.3) group. IFNAR1 functions in general as a heterodimer with IFNAR2, and, canonical signaling through the type I IFN receptor requires JAK1 and results in expression of interferon stimulated genes (ISGs), including IFI27 and PKR. Cell-surface IFNAR1 is a limiting factor for assembly of the functional type I IFN receptor complex. Intriguingly, IFNAR1 can form an active IFNB receptor by itself and activate signaling that does not involve activation of the Jak/STAT pathway (*32*). Among the transcripts uniquely upregulated by IFNAR1-IFNB signaling was TNFA, which in our study, together with IFNAR1, was increased in NC monocytes of participants in the TNFi non-response (IFNβ/α 0 or >1.3) group. The pattern of gene expression that differed between the patient groups could suggest that canonical type I IFN pathway signaling is increased in peripheral blood CL monocytes of RA patients who are likely to respond to TNFi, whereas Jak/STAT-independent IFNB-IFNAR1 signaling is increased in NC monocytes of those who are not likely to respond to TNFi. It is possible that this pattern could be exploited therapeutically in those who would be unlikely to respond to TNFi.

In summary, in this study we measured gene expression in single monocytes from seropositive RA patients prior to biologic treatment and compared data between groups defined by circulating IFNβ/α activity that would be predictive of TNFi response. The results suggest major differences in monocyte gene expression between the type I IFN activity groups, supporting downstream effects upon a critical effector cell population. Interestingly, JAK1 expression was a strong predictor of IFNβ/α activity group, and also was observed to be completely lacking in some patient’s monocytes. Our data suggest differences in IFNβ/α activity may skew canonical vs. non-canonical IFN pathway activation in RA patient monocytes.

## Data Availability

Data referred to in this manuscript is available upon reasonable request.

## 6 Author Contributions

TWM, WF, ZJ, and TN contributed to the conception and design of the study; TWM, WF, ZJ, MJ, JD, YGP, BD, DV, KW, TM, SP, CM, JD, and EM contributed to the acquisition of data; TWM, WF and TN contributed substantially to the analysis and interpretation of data; WF wrote the first draft of the manuscript; TWM and TN wrote subsequent drafts. All authors contributed to manuscript revision, read and approved the submitted version.

## References

1. L. Carmona, M. Cross, B. Williams, M. Lassere, L. March, Rheumatoid arthritis. Best Pract Res Clin Rheumatol 24, 733–745 (2010).

2. M. Boers, Understanding the window of opportunity concept in early rheumatoid arthritis. Arthritis Rheum 48, 1771–1774 (2003).

3. J. S. Smolen et al., Treating rheumatoid arthritis to target: recommendations of an international task force. Ann Rheum Dis 69, 631–637 (2010).

4. K. S. Upchurch, J. Kay, Evolution of treatment for rheumatoid arthritis. Rheumatology (Oxford) 51 Suppl 6, vi28–36 (2012).

5. J. M. Gwinnutt et al., Twenty-Year Outcome and Association Between Early Treatment and Mortality and Disability in an Inception Cohort of Patients With Rheumatoid Arthritis: Results From the Norfolk Arthritis Register. Arthritis Rheumatol 69, 1566–1575 (2017).

6. K. Puolakka et al., Early suppression of disease activity is essential for maintenance of work capacity in patients with recent-onset rheumatoid arthritis: five-year experience from the FIN-RACo trial. Arthritis Rheum 52, 36–41 (2005).

7. I. Filipovic, D. Walker, F. Forster, A. S. Curry, Quantifying the economic burden of productivity loss in rheumatoid arthritis. Rheumatology (Oxford) 50, 1083–1090 (2011).

8. T. Sokka et al., Work disability remains a major problem in rheumatoid arthritis in the 2000s: data from 32 countries in the QUEST-RA study. Arthritis Res Ther 12, R42 (2010).

9. C. M. Lopez de Padilla, T. B. Niewold, The type I interferons: Basic concepts and clinical relevance in immune-mediated inflammatory diseases. Gene 576, 14–21 (2016).

10. S. N. Kariuki et al., Genetic analysis of the pathogenic molecular sub-phenotype interferon-alpha identifies multiple novel loci involved in systemic lupus erythematosus. Genes Immun 16, 15–23 (2015).

11. T. Wampler Muskardin et al., Increased pretreatment serum IFN-beta/alpha ratio predicts non-response to tumour necrosis factor alpha inhibition in rheumatoid arthritis. Ann Rheum Dis 75, 1757–1762 (2016).

12. F. Liote, B. Boval-Boizard, D. Weill, D. Kuntz, J. L. Wautier, Blood monocyte activation in rheumatoid arthritis: increased monocyte adhesiveness, integrin expression, and cytokine release. Clin Exp Immunol 106, 13–19 (1996).

13. F. M. Batliwalla et al., Peripheral blood gene expression profiling in rheumatoid arthritis. Genes Immun 6, 388–397 (2005).

14. S. Sharma et al., Widely divergent transcriptional patterns between SLE patients of different ancestral backgrounds in sorted immune cell populations. J Autoimmun 60, 51–58 (2015).

15. D. Aletaha et al., 2010 rheumatoid arthritis classification criteria: an American College of Rheumatology/European League Against Rheumatism collaborative initiative. Ann Rheum Dis 69, 1580–1588 (2010).

16. T. B. Niewold, T. L. Rivera, J. P. Buyon, M. K. Crow, Serum type I interferon activity is dependent on maternal diagnosis in anti-SSA/Ro-positive mothers of children with neonatal lupus. Arthritis Rheum 58, 541–546 (2008).

17. K. A. Kirou et al., Coordinate overexpression of interferon-alpha-induced genes in systemic lupus erythematosus. Arthritis Rheum 50, 3958–3967 (2004).

18. C. E. Weckerle et al., Network analysis of associations between serum interferon-alpha activity, autoantibodies, and clinical features in systemic lupus erythematosus. Arthritis Rheum 63, 1044–1053 (2011).

19. T. B. Niewold, S. C. Wu, M. Smith, G. A. Morgan, L. M. Pachman, Familial aggregation of autoimmune disease in juvenile dermatomyositis. Pediatrics 127, e1239–1246 (2011).

20. X. Feng et al., Inhibition of interferon-beta responses in multiple sclerosis immune cells associated with high-dose statins. Arch Neurol 69, 1303–1309 (2012).

21. T. B. Niewold, J. Hua, T. J. Lehman, J. B. Harley, M. K. Crow, High serum IFN-alpha activity is a heritable risk factor for systemic lupus erythematosus. Genes Immun 8, 492–502 (2007).

22. Z. Jin et al., Single-cell gene expression patterns in lupus monocytes independently indicate disease activity, interferon and therapy. Lupus Sci Med 4, e000202 (2017).

23. H. Satomi, B. Wang, H. Fujisawa, F. Otsuka, Interferon-beta from melanoma cells suppresses the proliferations of melanoma cells in an autocrine manner. Cytokine 18, 108–115 (2002).

24. L. B. Ivashkiv, L. T. Donlin, Regulation of type I interferon responses. Nat Rev Immunol 14, 36–49 (2014).

25. K. Fink et al., IFNbeta/TNFalpha synergism induces a non-canonical STAT2/IRF9-dependent pathway triggering a novel DUOX2 NADPH oxidase-mediated airway antiviral response. Cell Res 23, 673–690 (2013).

26. S. J. Rodig et al., Disruption of the Jak1 gene demonstrates obligatory and nonredundant roles of the Jaks in cytokine-induced biologic responses. Cell 93, 373–383 (1998).

27. T. Ito, H. Kanzler, O. Duramad, W. Cao, Y. J. Liu, Specialization, kinetics, and repertoire of type 1 interferon responses by human plasmacytoid predendritic cells. Blood 107, 2423–2431 (2006).

28. A. Prakash, E. Smith, C. K. Lee, D. E. Levy, Tissue-specific positive feedback requirements for production of type I interferon following virus infection. J Biol Chem 280, 18651–18657 (2005).

29. M. Severa et al., Sensitization to TLR7 agonist in IFN-beta-preactivated dendritic cells. J Immunol 178, 6208–6216 (2007).

30. U. Nir, L. Maroteaux, B. Cohen, I. Mory, Priming affects the transcription rate of human interferon-beta 1 gene. J Biol Chem 260, 14242–14247 (1985).

31. G. Weiss et al., MyD88 drives the IFN-beta response to Lactobacillus acidophilus in dendritic cells through a mechanism involving IRF1, IRF3, and IRF7. J Immunol 189, 2860–2868 (2012).

32. N. A. de Weerd et al., Structural basis of a unique interferon-beta signaling axis mediated via the receptor IFNAR1. Nat Immunol 14, 901–907 (2013).

